# Estimating the effect of mobility on SARS-CoV-2 transmission during the first and second wave of the COVID-19 epidemic in Switzerland: a population-based study

**DOI:** 10.1101/2021.04.16.21255636

**Authors:** Adrian Lison, Joel Persson, Nicolas Banholzer, Stefan Feuerriegel

**Affiliations:** ETH Zurich, 8092 Zurich, Switzerland

## Abstract

The effect of mobility and its value for surveillance in different waves of the COVID-19 epidemic is still unclear. In this study, we compared the role of mobility during the first and second epidemic wave in Switzerland by analysing the link between daily travel distances and the effective reproduction number *R*_*t*_ of SARS-CoV-2. Here we used aggregated mobile phone data from a representative panel survey of the Swiss population to measure human mobility. We estimated the effects of reductions in daily travel distance on *R*_*t*_ via a regression model. We compared mobility effects between the first wave (March 2-April 7, 2020) and the second wave (October 1-December 10, 2020) across mode of transport, travel purpose, sociodemographic subgroup and movement radius. We found that human mobility was associated with the effective reproduction number of SARS-CoV-2 during both the first and second epidemic wave in Switzerland. The estimated relative effects of mobility were similar in both waves for all modes of transport, travel purposes, and sociodemographic subgroups but differed by movement radius. Moreover, smaller mobility reductions in the second wave translated into smaller overall reductions of *R*_*t*_. Mobility data from mobile phones have a continued potential to support real-time surveillance of COVID-19 during epidemic waves.

## 1. Introduction

The global epidemic of COVID-19 is ongoing, with repeated surges in community transmission across many European countries.[1] For instance, in Switzerland, the effective reproduction number of SARS-CoV-2 surpassed the threshold of one during both a first wave in spring 2020 and a second wave in fall 2020. To enable timely surveillance of the epidemic, the use of mobile phone data has been proposed.[2–4] Mobile phone data can capture human movements in near real-time and thus serve as a proxy for population-level mobility under COVID-19 policy measures.[5–7] Initially, such mobility data have been used to understand the spreading of COVID-19 out of Wuhan, China.[8, 9] Moreover, the role of mobility in different countries has been studied for the first wave of the epidemic. For instance, some studies found a link between disease spreading and the frequency or duration of visits at specific points of interest (e. g., parks, shops, restaurants).[10–12] Others have connected human movements, measured for example through travel distances, to the course of the first wave.[13–17] On the other hand, for time periods after the first wave, some studies have suspected a decoupling of the relationship between mobility and disease transmission.[18–20] However, to the best of our knowledge, no study to date has used real-world data to compare the effects of mobility in different epidemic waves. The specific role of mobility and its value for surveillance during later waves of the COVID-19 epidemic has thus remained largely unclear.

In this study, our aim was to compare the role of mobility during the first and the second epidemic wave of COVID-19 in Switzerland. Our analysis focused on epidemic waves, because we expect population-level behaviour to be most relevant for disease control under large-scale community transmission. The study periods were defined as March 2–April 7, 2020, i. e., the first wave, and October 1–December 10, 2020, i. e., the second wave (see Section 2 for definition). For each wave, we estimated the effect of human mobility on the effective reproduction number of SARS-CoV-2. Here, mobility was measured by daily travel distances based on aggregated mobile phone data from a representative, longitudinal panel survey (*N* = 2561 participants). Formally, we linked mobility to the effective reproduction number using a regression model and then compared the estimated mobility effects between the first and second wave for the study population. We further analysed how the role of mobility by mode of transport, travel purpose, and sociodemographic subgroup (age group, household size, employment status, and urbanization level) differed between the waves. We also estimated how the movement radius was linked to the effective reproduction number.

## 2. Methods

### Study periods

The study periods were selected based on criteria for disease control from the World Health Organization.[21] Specifically, the selected study periods had both a high average number of transmissions per infectious person (indicated by an effective reproduction number above 1) and a substantial proportion of infectious persons (indicated by a test positivity rate above 5 %) in the previous three weeks. For Switzerland, this resulted in two study periods from March 2– April 7, 2020, and from October 1–December 10, 2020, hereafter referred to as first and second wave. The end of the second study period was set to two weeks before Christmas Eve to exclude changes in reporting practice during the Christmas holidays.

### Mobility data

We measured human mobility using data from a mobile phone-based panel survey. The panel survey was conducted by intervista AG on behalf of the Swiss National COVID-19 Science Task Force. It included *N* = 2561 participants that were representative of the population in Switzerland by age, gender, and region.[22] Moreover, sociodemographic characteristics of each participant were recorded: age group (15–29, 30–64, or 65–79 years), household size (1, 2, or *>*3 persons), employment status (employed, unemployed, or in education), and urbanization level for the place of residence (rural or urban area). Daily releases of the data were made available throughout the epidemic.

Movements of each participant were continuously tracked throughout the year 2020 via triangulation between cell towers and Wi-Fi hotspots, data from movement sensors of the phone, and interactions with Bluetooth beacons.[22] Based on the movement data, the absolute daily travel distance and the movement radius (both in km) were computed for each participant and aggregated over the study population. Analogous to the estimates of the effective reproduction number, the aggregates were smoothed with a 3-day moving average. The movement radius is defined as the maximum distance from place of residence on a given day. The recorded travel distance was further stratified by mode of transport (car/motorcycle, public transport, foot, or other) and travel purpose (occupation, shopping, or leisure). Details are provided in Supplement A.

### Effective reproduction number and policy measures

The effective reproduction number *R*_*t*_ denotes the expected number of secondary infections resulting from an infection at time *t* and is used to monitor disease transmission over time. Estimates of *R*_*t*_ in Switzerland were obtained from the Swiss National COVID-19 Science Task Force.[23, 24] The underlying estimation procedure was based on the time series of newly hospitalized patients with COVID-19 and adjusted for the incubation period and time between symptom onset and hospitalization. The estimates were provided as a 3-day moving average. We used point estimates of the effective reproduction number for our analysis. In the descriptive plots, we also report uncertainty intervals.

Relevant COVID-19 policy measures in Switzerland were selected using a systematic procedure. Implementation dates of the policy measures were obtained from the official regulations of the Swiss Federal Council and checked against dates from the Oxford Government Response Tracker[25] and the Swiss National COVID-19 Science Task Force[23]. Details on the systematic procedure and encoding of policy measures are provided in Supplement A. For the first wave, the dates of policy measures were encoded as March 13, 2020 (ban on public gatherings of over 100 people), March 17, 2020 (closures of schools and venues), and March 21, 2020 (ban on public gatherings of over 5 people). For the second wave, the dates of policy measures were encoded as October 19, 2020 (ban on public gatherings of over 15 people) and October 28, 2020 (venue restrictions and ban on private meetings of over 10 people). Each of the aforementioned measures were implemented nationwide (i. e., across all cantons in Switzerland).

### Statistical analysis

We linked mobility to the effective reproduction number of SARS-CoV-2 using a regression model. As in earlier research,[26] the effective reproduction number *R*_*t*_ on day *t* was assumed to follow a gamma distribution. We then modelled a log-linear relationship between the expected value of *R*_*t*_ and the observed average daily travel distance on the same day. We further applied a logarithmic transformation such that changes in mobility could be measured on a relative scale. We accounted for the effects of policy measures by adding dummy variables indicating whether a measure was in effect on day *t* or not. For each weekday, a different intercept was included to capture potential differences between weekdays in reporting and mobility (e. g., due to less reporting and mobility on weekends). In the resulting model, the coefficient of mobility can be interpreted as the expected percentage change in *R*_*t*_ associated with a 1 % change in mobility, conditional on the policy measures and the day of the week.

We further analysed how the effect of mobility by mode of transport, travel purpose, and sociodemographic subgroup (age group, household size, employment status, and urbanization level) differed between the waves. A separate model was fitted for each mode of transport, for each travel purpose, and for each sociodemographic subgroup, with the stratified average daily travel distance as explanatory variable. All other variables were analogous to the main model from above.

We also analysed the extent to which the movement radius was linked to the effective reproduction number. The movement radius was categorized into residential (*<*500 m), local (500 m– 2 km), municipal (2 km–10 km), regional (10–50 km), or long-range (*>*50 km) mobility. Then, the daily share of the study population travelling within each movement radius was computed, and, for each movement radius, a separate regression model was fitted where the aforementioned share was the explanatory variable. Here, the coefficient of movement radius can be interpreted as the expected percentage change in *R*_*t*_ associated with a one percentage point increase in the share of mobility within a certain movement radius. Otherwise, the model had the same specification as the model based on travel distance.

The specifications of the different models are provided in Supplement B. The models were fitted separately for the first wave (using data from March 2–April 7, 2020) and the second wave (using data from October 1–December 10, 2020). This enabled us to estimate the effects of mobility independently for both waves, while respecting potential differences in epidemiological features or reporting practices.

All model parameters were estimated in a fully Bayesian framework. Estimation was conducted in R 4.0.3 and Stan 2.01.0 using Markov chain Monte Carlo (MCMC) sampling via the No-U-Turn sampler (NUTS). Four chains were run, with 1000 warm-up iterations and 1000 sampling iterations each. We defined weakly informative priors for all model parameters (see Supplement B). The estimates were checked using common Bayesian model diagnostics, and indicated good model fit, sufficient effective sample size, convergence of the chains, and absence of particularly influential observations (see Supplement C). Unless stated otherwise, we report the posterior mean and the 95 % credible interval (CrI) of estimated parameters.

As part of our robustness checks, we tested alternative estimates of the effective reproduction number and other study periods. We also extended our model to account for potential changes in testing intensity. This led to qualitatively similar findings.

### Ethical statement

Movement data from the panel survey were collected in anonymized form in line with the Federal Act on Data Protection and the General Data Protection Regulation. All participants consented to the use for scientific purposes. Only aggregated data were analysed in this study. Ethics approval for the study was obtained by the institutional review board at ETH Zürich.

## 3. Results

The effective reproduction number *R*_*t*_ is shown in Fig. 1 for both study periods: March 2– April 7, 2020, i. e., the first wave, and October 1–December 10, 2020, i. e., the second wave, in Switzerland. During both waves, *R*_*t*_ was initially above the threshold of one (indicating exponential growth) but decreased over time. In the first wave, *R*_*t*_ fell below the threshold of one from March 18, 2020 onward. In the second wave, *R*_*t*_ first fell temporarily below the threshold of one on October 26, 2020 but surpassed the threshold again on November 23, 2020.

**Figure 1:**
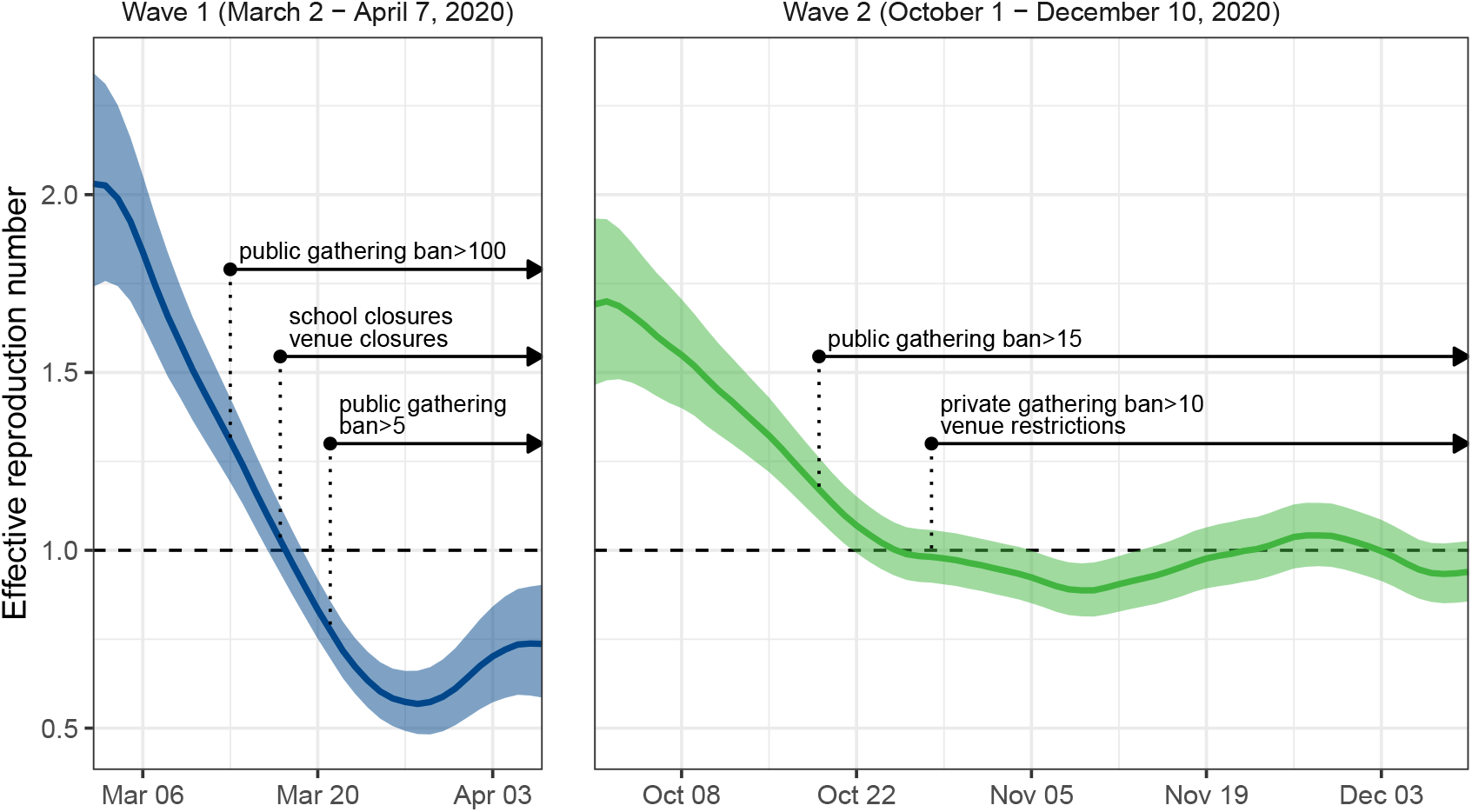
Effective reproduction number during the first and second wave in Switzerland. Shown is the effective reproduction number for the first wave (March 2–April 7, 2020) and second wave (October 1– December 10, 2020) in Switzerland. Shaded areas indicate the 95 % uncertainty interval. Annotations show the implementation dates of policy measures. Once introduced, policy measures remained in effect until the end of both study periods.

### Changes in mobility

We measured relative reductions in mobility during the epidemic as percentage change from the average daily travel distance in February, 2020. As shown in Fig. 2A, mobility decreased considerably in both waves. In the first wave, a large reduction in daily travel distances occurred from March 13, 2020 onward. The strongest change was reached on March 29, 2020, with a reduction of 73*·*37 %. In the time period between the first and second wave, mobility increased, and, as a result, daily travel distances were of a similar magnitude during July–October as in February. In the second wave, the reduction in daily travel distances was more gradual and of smaller size. Here, the strongest change was reached on December 6, 2020 with a reduction of 43*·*63 %.

**Figure 2:**
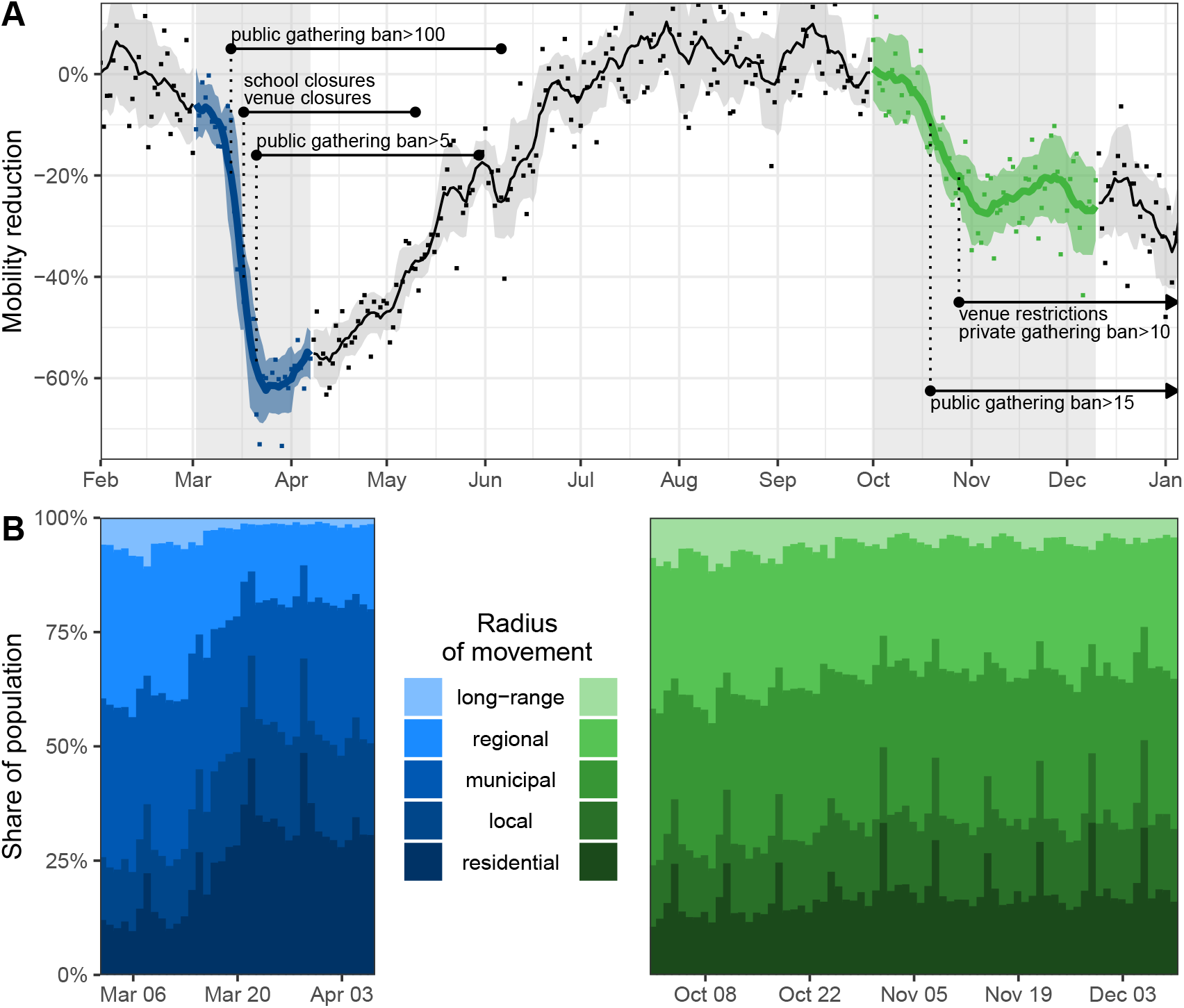
Human mobility in Switzerland. (A) Shown are reductions in average daily travel distance relative to the February average. Dots represent daily reductions and shaded trend lines show the corresponding 7-day moving average with standard deviation. Annotations show the implementation dates of policy measures. (B) Shown is the share of the population travelling within a certain movement radius.

The share of mobility within a smaller movement radius increased considerably in the first wave and slightly in the second wave (Fig. 2B). For example, the share of the population traveling within a residential radius (*<*500 m) was at most 28*·*50 % in February, whereas it reached up to 48*·*29 % (March 19, 2020) in the first wave and up to 33*·*00 % (November 29, 2020) in the second wave. In contrast, the share of the population traveling within a long-range radius (*>*50 km) decreased. The share accounted for up to 6*·*03 % of the study population in February, but the share declined to 1*·*15 % (March 31, 2020) in the first wave and 3*·*59 % (November 4, 2020) in the second wave.

Relative mobility reductions varied notably across modes of transport, with walking affected least and public transport affected most (Fig. 3A). As such, daily travel distances by public transport reached a maximum reduction of 87*·*23 % (March 29, 2020) in the first wave and 56*·*72 % (November 6, 2020) in the second wave. They were also lower than the average in February, 2020 during the time between the waves. Daily travel distances by car/motorcycle decreased considerably in the first wave with a maximum reduction of 70*·*94 % (March 22, 2020), but less strongly in the second wave with a maximum reduction of 37*·*09 % (December 6, 2020). Across different travel purposes, mobility reductions were mostly similar in both waves (Fig. 3B). Across different sociodemographic subgroups, mobility also showed broadly similar declines. This was observed across all age groups, employment statuses, household sizes, and urbanization levels (Fig. 3C-F).

**Figure 3:**
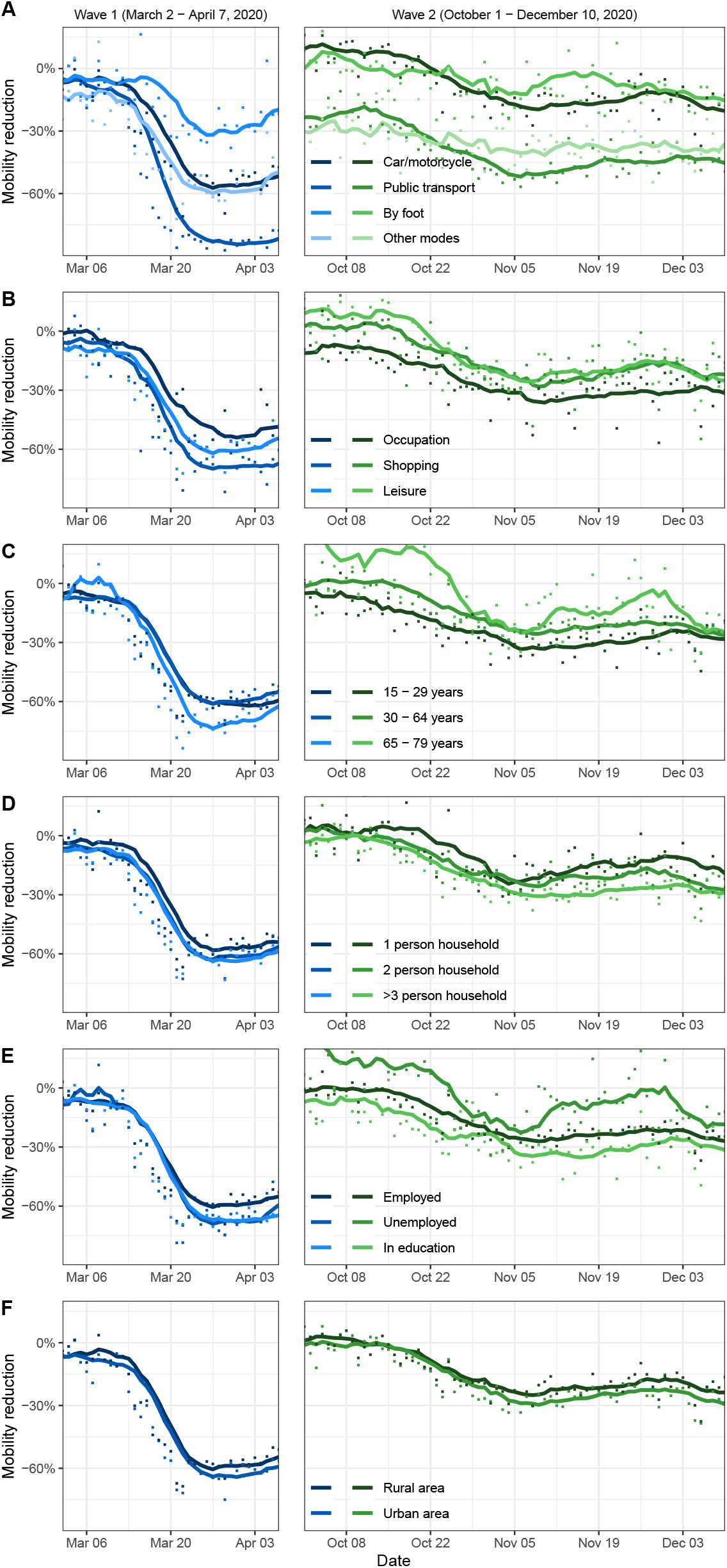
Comparison of relative mobility reductions over time. For both waves, reductions in average daily travel distance, relative to the February average, are shown. Dots represent daily reductions and trend lines show the corresponding 7-day moving average. Comparisons are shown for (A) mode of transport, (B) travel purpose, (C) age group, (D) household size, (E) employment status, and (F) urbanization level.

### Estimated effects of mobility

The effect of mobility on the effective reproduction number *R*_*t*_ was estimated separately for both waves using our regression model. The estimated coefficients for the effect of mobility were of similar magnitude in the first and second wave. As shown in Fig. 4A, the estimated reduction in *R*_*t*_ per 1 % reduction in average travel distance of the population was 0*·*73 % (95 % CrI 0*·*34–1*·*03 %) in the first wave and 1*·*04 % (95 % CrI 0*·*66–1*·*42 %) in the second wave. Similarities in the relative effect of mobility between the first and second wave were found across all modes of transport, travel purposes, and sociodemographic subgroups (age group, household size, employment status, and urbanization level). In all cases, the 80 % credible intervals for mobility effects overlapped between the first and second wave (Fig. 4B-G).

**Figure 4:**
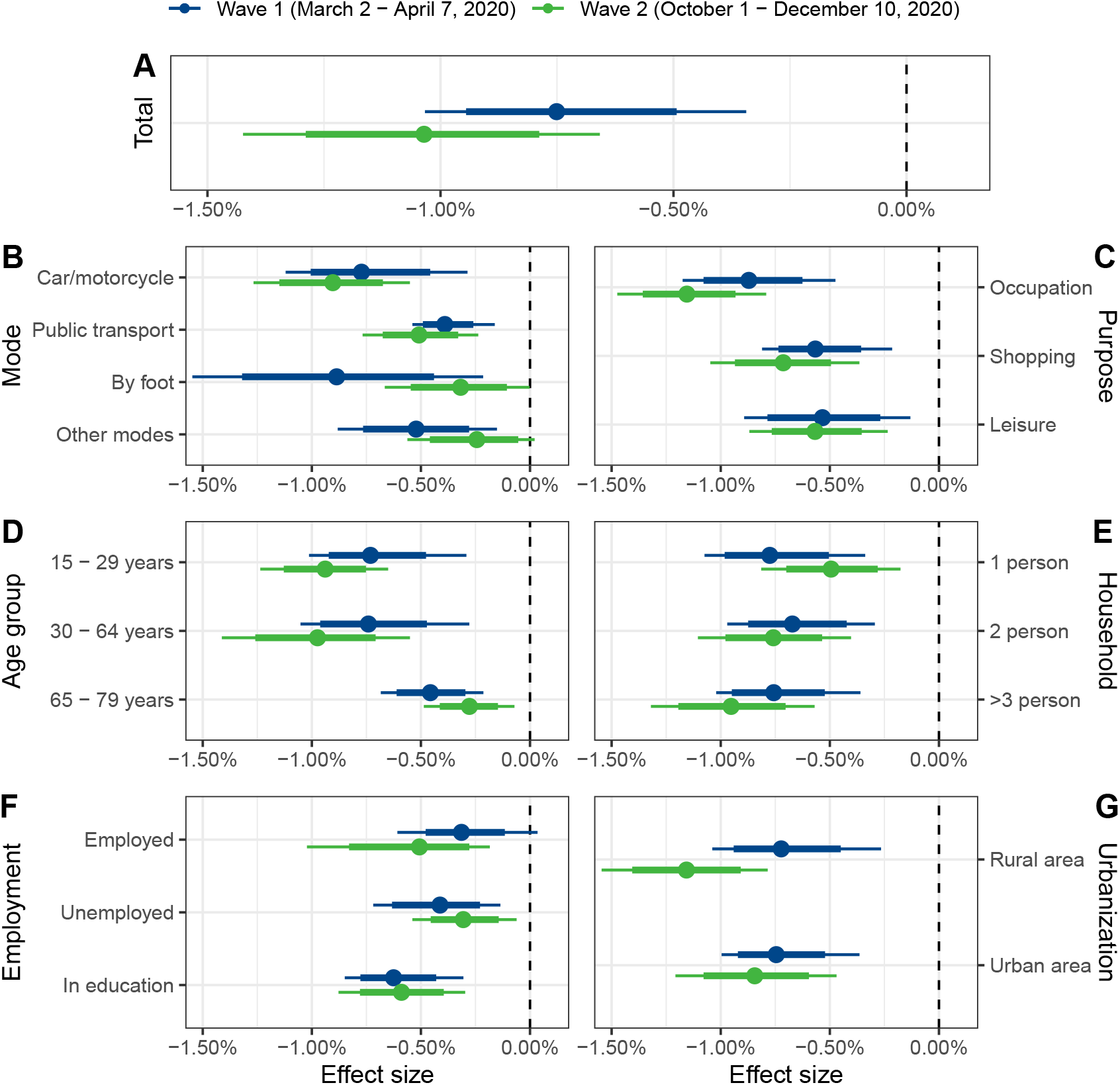
Estimated relative effects of mobility. Shown are the estimated changes in the effective reproduction number given a 1 % reduction in mobility, for the first and second wave, respectively. Comparisons are shown for (A) the total study population, and across (B) mode of transport, (C) travel purpose, (D) age group, (E) household size, (F) employment status, and (G) urbanization level. Posterior means (dots) and the 80 % and 95 % credible intervals (thick and thin bars) are reported.

Figure 5 shows the estimated overall effect of observed mobility reductions during the first and second wave. As the observed reduction in mobility was larger for the first wave than the second wave, the overall change in the effective reproduction number was also larger for the first wave (even though the estimated relative mobility effects were similar). In the first wave, the maximum overall reduction in *R*_*t*_ attributed to mobility by the model was estimated to be 51*·*56 % (95 % CrI 28*·*30–65*·*28 %). Together with the implemented policy measures, this corresponds to an estimated effective reproduction number of 0*·*61 (95 % CrI 0*·*47–0*·*78), which is below the threshold of one with high probability. In the second wave, the maximum overall reduction was estimated to be 31*·*94 % (95 % CrI 21*·*27–41*·*44 %). Together with the implemented policy measures, this corresponds to an effective reproduction number of 0*·*87 (95 % CrI 0*·*75– 1*·*00), which is not below the threshold of one with high probability. Hence, the comparatively smaller mobility reductions in the second wave also translated into smaller reductions of the effective reproduction number.

**Figure 5:**
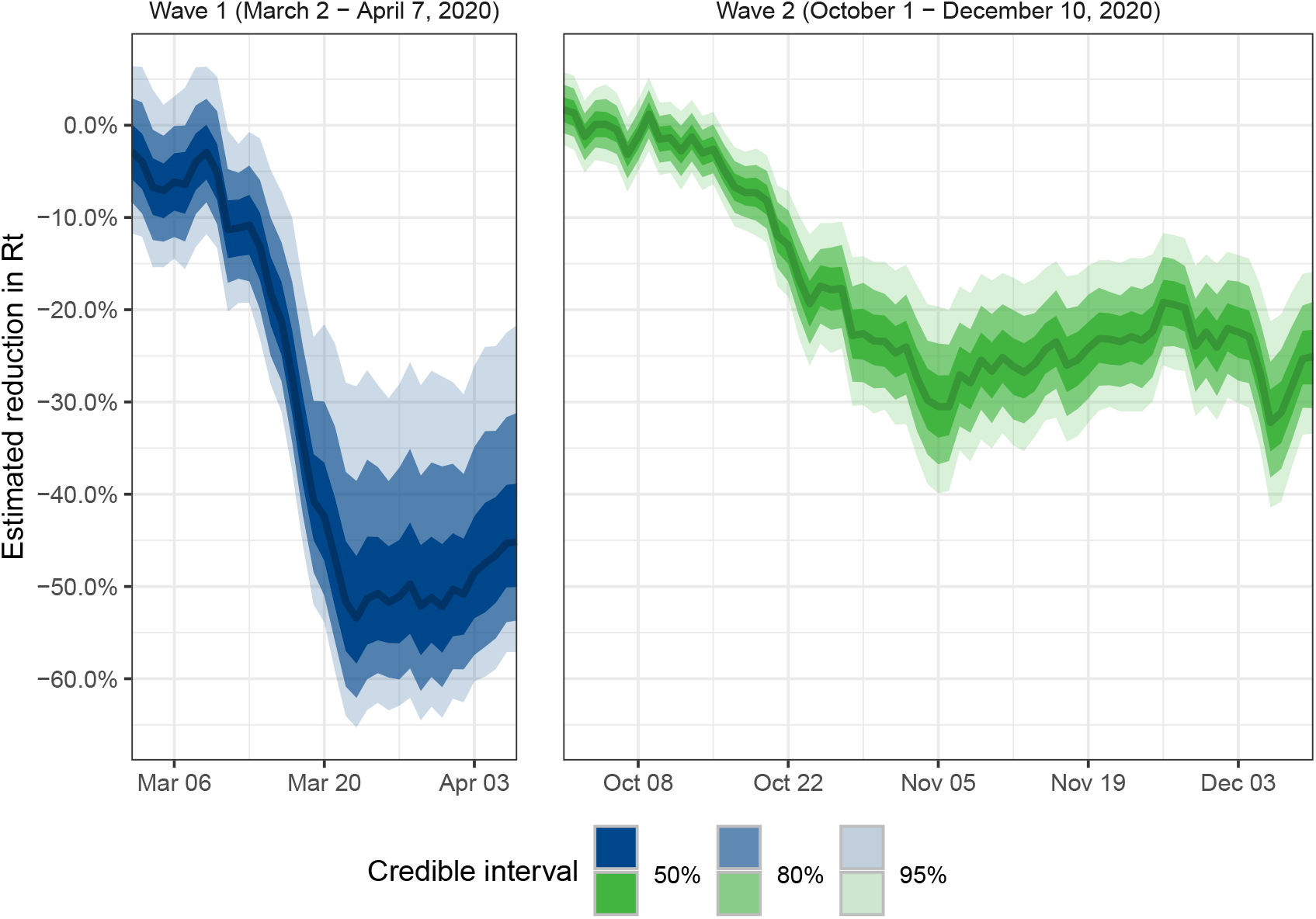
Estimated overall effect of mobility. Shown are the daily estimated reductions in the effective reproduction number attributed to mobility by the model in the first and second wave. Posterior means (lines) and 50 %, 80 % and 95 % credible intervals (shaded areas) of *R*_*t*_ are reported.

We furthermore analysed the extent to which the movement radius was linked to the effective reproduction number *R*_*t*_ (Fig. 6). The estimated effects of residential, local, regional, and long-range movement radius were similar in both waves, with the corresponding 80 % credible intervals for the two waves overlapping. The effective reproduction number *R*_*t*_ was negatively associated with the residential movement radius and positively associated with both the regional and the long-range movement radius in both waves. The local movement radius only had a significant association with *R*_*t*_ in the second wave, where the estimated change in *R*_*t*_ per one percentage point reduction in the population share was 5*·*31 % (95 % CrI 2*·*00–8*·*32 %). Moreover, the effect of a municipal movement radius likely differed between the two waves. In the first wave, we estimated a −2*·*71 % (95 % CrI −5*·*59–0*·*20 %) change in *R*_*t*_ per one percentage point reduction in the corresponding population share. In the second wave, the same effect was estimated to be 1*·*61 % (95 % CrI −1*·*94–5*·*14 %).

**Figure 6:**
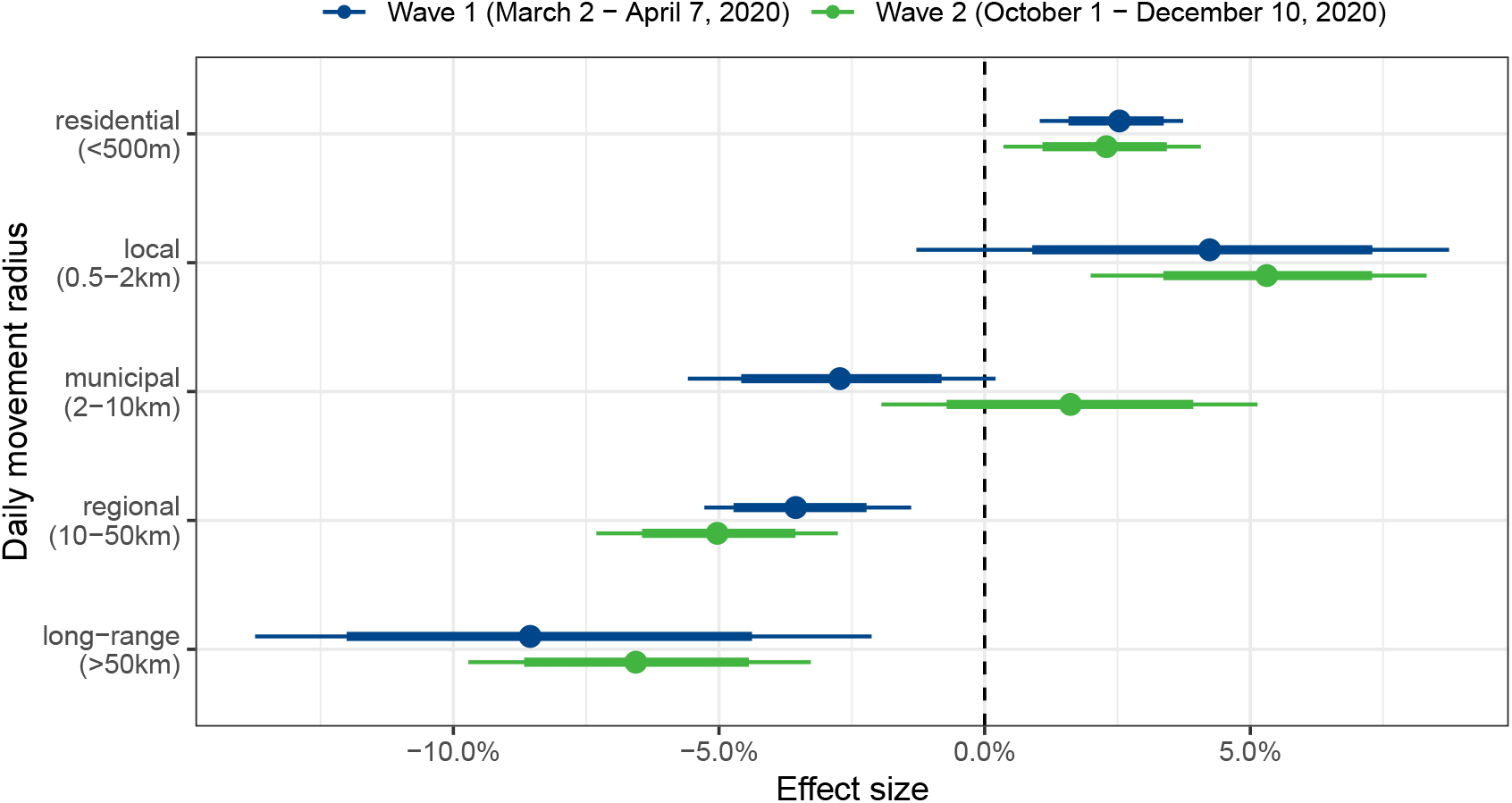
Estimated mobility effects across movement radius. Shown are the estimated changes in *R*_*t*_ given a one percentage point decrease in the share of the population traveling within a certain movement radius. Posterior means (dots) and the 80 % and 95 % credible intervals (thick and thin bars) are reported.

### Robustness checks

To assess the robustness of our results, we tested other study periods for defining the first and second wave, other estimates of the effective reproduction number, an extended model with autocorrelated error terms, and a model adjusting for the effect of testing intensity (see Supplement D). In all tests, the estimated effects of mobility for both waves were robust. In addition, we found no significant effect of mobility when fitting our model for the time period in between the first and the second wave, which generally corresponds to a phase with fairly low case numbers.

## 4. Discussion

In this work, we studied how mobility was linked to the effective reproduction number of SARS-CoV-2 in Switzerland, a country where travel for work, education, and leisure is widespread.[27] For this, mobility data were obtained from a mobile phone-based panel survey, whose participants were representative of the population by age, gender and region. Our research on the link between mobility and COVID-19 spreading extends beyond previous studies [10–18] by comparing the effect of mobility in two different waves.

As our results show, mobility in Switzerland reduced considerably during the first wave (defined as March 2–April 7, 2020) and to a lesser extent in the second wave (October 1–December 10, 2020). This observation can reflect differences in population behaviour and the policies adopted by the Swiss government. In particular, while venue closures led to an overall shutdown in the first wave, policies in the second wave initially focused on contact tracing and hygiene measures with fewer restrictions on public life,[28] a pattern which has been observed in many European countries.[25] We furthermore found similar relative reductions in mobility across travel purposes and sociodemographic subgroups, implying mostly homogeneous mobility changes among the population in Switzerland. Differences in mobility reduction were observed between modes of transport, indicating a tendency of the population to avoid public transport.

Using our regression model, we linked reductions in mobility to changes in the effective reproduction number of SARS-CoV-2, adjusting for policy measures. Previous work found a positive association between mobility and transmission in the first wave but it was suspected for many countries worldwide that a decoupling in the relationship took place afterwards.[18–20] Our results suggest that human mobility is an important determinant for explaining reductions in SARS-CoV-2 transmission during both the first and the second wave in Switzerland. We found that the effect of a 1 % reduction in average daily travel distance on the effective reproduction number was of similar magnitude in both waves. This result was consistent across modes of transport, travel purposes, and sociodemographic subgroups. Our findings therefore indicate that, during epidemic waves, population-level measures of mobility can explain changes in transmission.

Differences between the two waves were found regarding the overall effect of mobility, since smaller mobility reductions in the second wave also translated into smaller overall reductions of the effective reproduction number. Furthermore, our analysis of the link between movement radius and effective reproduction number suggests a more pronounced role of stay-at-home behaviour during the first wave, in which a larger movement radius was more strongly associated with increases in *R*_*t*_.

This study is subject to several limitations. First, our analysis was limited to Switzerland. While the overall course of the epidemic in Switzerland is characterized by a similar variability seen in many European countries, a future comparison with other countries could be especially valuable to investigate country-specific differences in mobility behaviour. Second, we used mobility data only from a sample of the population in Switzerland, which was however selected to be representative by age, gender, and region. Third, in our study, mobility was measured via the daily travel distance. Other studies have analysed mobility in the first wave using alternative metrics such as the radius of gyration or trip counts,[2] which may have different interpretations. However, as our model measures changes in mobility on a relative scale, we expect our results not to be highly sensitive to the choice of metric.

In our models, both the mobility variables and the time-varying effective reproduction number *R*_*t*_ were smoothed with a 3-day moving average. Our estimates are therefore with respect to smoothed mobility and *R*_*t*_. We further acknowledge that the estimation of *R*_*t*_ can entail considerable uncertainty, especially when the number of reported cases is low. We addressed this limitation by selecting only study periods with a substantial proportion of infectious persons and thus comparatively small uncertainty. To capture structural changes in transmission aside from mobility, we fitted separate models for both the first and second wave and included variables for policy measures and weekday-specific intercepts. Still, it cannot be ruled out that there are further factors not included in our model that could explain the changes in mobility and the effective reproduction number. Our estimates should hence be interpreted as providing associative and not causal relationships. Lastly, it is important to note that our study estimated the overall effect of mobility on the effective reproduction number. We thereby did not model the effects of other behavioural changes such as mask-wearing, which could have moderated the effect of mobility.

## 5. Conclusion

Our study highlights the continued value of mobile phone data in the context of real-time disease surveillance of COVID-19. The observed link between mobility and epidemic spreading was already found for other diseases.[29, 30] Here, we provide evidence that changes in the time-varying effective reproduction number, which becomes available only with a time lag of several days or weeks, can be predicted by mobility data available on the same day. Thus, digital tracking of human movements may provide an opportunity for real-time assessment of the epidemic situation, ahead of traditional reporting. For this purpose, mobile phone-based panel surveys can provide representative mobility data while ensuring consent and privacy protection. Mobility data can be further stratified by movement radius, mode of transport, travel purpose, or sociodemographic subgroups, to enable further insights for disease surveillance. Such insights can be made available in real time, for example in the form of social-distancing indicators. This can contribute to timely and informed policy-making during the COVID-19 epidemic.

## Data Availability

All data used in this study are publicly available. Mobility data are available from intervista AG. Estimates of the effective reproduction number are available from the Swiss National COVID-19 Science Task Force. Data on policy measures and testing are available from the Swiss Federal Office of Public Health.

https://www.intervista.ch/media/2020/03/Report_Mobilit%C3%A4ts-Monitoring_Covid-19.pdf

https://sciencetaskforce.ch/en/current-situation/

https://www.bag.admin.ch/bag/en/home.html

## Ethics approval

Ethics approval (EK 2020-N-179) for this study was obtained by the institutional review board (“Ethics Commission”) at ETH Zurich.

## Declaration of interests

SF declares membership in a COVID-19 Working Group by the World Health Organization (WHO) but without competing interests. SF reports grants from the Swiss National Science Foundation outside of the submitted work. All other authors declare no competing interests.

## Funding

This work has received financial support from the Swiss National Science Foundation (SNSF) as part of the Eccellenza grant 186932 on “Data-driven health management”. The funding source had no role in study design, data collection, data analysis, interpretation, writing of the manuscript, or decision to submit. All authors had full access to all the data used in the study and accept responsibility to submit for publication.

## Data sharing

All data used in this study are publicly available. Mobility data are available from intervista AG.[22] Estimates of the effective reproduction number are available from the Swiss National COVID-19 Science Task Force.[23] Data on policy measures and testing are available from the Swiss Federal Office of Public Health.

## Notes

### Author Declarations

Ethics approval (EK 2020-N-179) for this study was obtained by the institutional review board at ETH Zurich.

