## Supplementary material for "Estimating the effect of mobility on SARS-CoV-2 transmission during the first and second wave of the COVID-19 epidemic in Switzerland: a population-based study"

### A. Data

#### *A.1. Mobility data*

The mobility survey in Switzerland during COVID-19 was conducted by the company intervista AG on behalf of the Swiss Federal Statistical Office, the Statistical Office of the Canton of Zurich, the Swiss National COVID-19 Science Task Force, and the KOF Swiss Economic Institute.[22] The study population was chosen to be representative by age, gender, and region, according to targets provided by the Swiss Federal Statistical Office. Data were collected in anonymized form in line with the Federal Act on Data Protection and the General Data Protection Regulation. All participants consented to the use for scientific purposes. Ethics approval for this study was obtained by the institutional review board at ETH Zürich.

Data on the daily travel distance and movement radius of the participants were provided in aggregated form (mean and interval-censored distribution). Analogous to the estimates of the effective reproduction number, the data were smoothed with a 3-day moving average. Moreover, the data were stratified by mode of transport, travel purpose, and various sociodemographic characteristics. Mode of transport and travel purpose were inferred from probabilistic models. For this, data on location and speed, sensor data from the phone, site data (e. g., public transport stations, shops, restaurants), and profiles of participants (e. g., car ownership) were used. Modes of transport were categorized into "car/motorcycle", "public transport", "foot", and "other modes". Here, "public transport" refers to travel by train, tram, or bus, and "other modes" refers to travel by bike, mountain railway, or ship. Travel purposes were categorized into "occupation", "shopping" and "leisure". Here, "occupation" refers to work and education, and "leisure" refers to recreational activities, including restaurant visits. The movement radius was measured as the maximum distance from the place of residence.

Sociodemographic characteristics, i. e., age group, employment status, household size, and urbanization level, were obtained from digital polls. The urbanization level was inferred from the place of residence and categorized into "urban" and "rural", according to criteria of the Swiss Federal Statistical Office. Here, "urban areas" were defined as municipal centres and agglomeration, while "rural areas" were defined as countryside and suburban municipalities.

#### *A.2. Effective reproduction number*

Estimates of the time-varying effective reproduction number $R_t$ of COVID-19 in Switzerland were provided by the Swiss National COVID-19 Science Task Force.[23] The estimates are based on publicly available data and additional information from the Federal Office of Public Health.[24] The delay between infection and reporting of cases was taken into account. Hence, the estimates of $R_t$ on day $t$ represent the average transmission per person on that respective day $t$, but can as such only be obtained with a delay of several weeks. The estimates were provided as a 3-day moving average.

#### *A.3. Policy measures*

The policy measures considered in this study were selected in a three-step procedure as follows. First, we collected all national COVID-19 policy measures in Switzerland and their corresponding implementation dates from the official regulations of the Swiss Federal Council. Second, we filtered for policy measures that have been considered as relatively influential by recent studies,[4, 5] that is, restrictions concerning private or public gatherings, businesses, or educational facilities. Third, we checked the dates of the policy measures obtained against the dates from the Oxford Government Response Tracker and the Swiss National COVID-19 Science Task Force and found them to be in agreement.

Table A.1 lists the policy measures considered in this study. Note that the measures were each implemented nationwide on the same date. Furthermore, school closures were implemented at both the primary and the secondary level.

In the first wave, school closures and venue closures were implemented with a time difference of one day. To ensure identifiability of their effects, both measures were combined by encoding them as a single dummy variable active from March 17, 2020 onward. Our model diagnostics show that this does not substantially influence the estimates. In the second wave, contact restrictions and early closing hours came into effect on the same day. Hence, they were also encoded together as a single dummy variable.

| Measure | Description | Time period |
|---|---|---|
| Public gathering ban >100 | Ban on gatherings of more than 100 people at events | 13-03-2020 – 06-06-2020 |
| School closure | Closure of all schools, higher education, and other educational facilities | 16-03-2020 – 10-05-2020 |
| Venue closure | Closure of all shops, restaurants, bars, and recreational facilities | 17-03-2020 – 10-05-2020 |
| Public gathering ban >5 | Ban on gatherings of more than 5 people in public places | 21-03-2020 – 30-05-2020 |
| Public gathering ban >15 | Ban on gatherings of more than 15 people in public places | 19-10-2020 – end of study |
| Private gathering ban >10 | Ban on private meetings of more than 10 people | 28-10-2020 – end of study |
| Venue restrictions | Occupancy limits, closing hours from 23 pm – 6 am for restaurants & bars, closure of night clubs | 28-10-2020 – end of study |

**Table A.1:** Overview of policy measures in this study.

### B. Statistical analysis

#### *B.1. Study periods*

The choice of study periods was based on criteria by the World Health Organization[21] on disease control. Time periods with both (1) a high average number of transmissions per infectious person and (2) a substantial proportion of infectious persons in the previous three weeks were selected as follows. (1) A high average number of transmissions per infectious person was defined by the effective reproduction number being above the threshold of one. (2) A substantial proportion of infectious persons was defined by the share of positive COVID-19 tests (test positivity rate) being above 5 %. Data on testing in Switzerland was obtained from the Federal Office of Public Health. We used a 3-day moving average to smooth out weekday effects in testing.

The above criteria returned two time periods: March 2–April 7, 2020 (first wave) and October 1–December 10, 2020 (second wave). The end of the second period was set two weeks before Christmas Eve to exclude changes in reporting practice during the Christmas holidays.

*B.2. Main model*

We linked mobility to the effective reproduction number of COVID-19 via a Bayesian regression model. The effective reproduction number on day $t$, denoted by $R_t$, was modelled to follow a gamma distribution, i. e.,

$$R_t \sim \text{Gamma}(\alpha, \beta_t)\,. \tag{B.1}$$

Here, $\alpha$ and $\beta_t$ is the shape and rate parameter of the gamma distribution, respectively, that define the expected value of the effective reproduction number on a given day $t$ as $\mathbb{E}[R_t] = \mu = \frac{\alpha}{\beta_t}$. We modelled the expected value with the following regression function of the explanatory variables using a log-link:

$$\mu(R_t) = \exp(\delta_{\text{wd}(t)} + \sum_{j=1}^{J} \gamma_j p_{j,t} + \theta \log \bar{m}_t)\,. \tag{B.2}$$

The term $\delta_{\text{wd}(t)}$ is an intercept allowed to vary by weekday in order to capture differences between weekdays in reporting and mobility (e. g., due to less reporting and mobility on weekends) that could confound the relationship between mobility and the effective reproduction number. To control for confounding by policy measures, the dummy variable $p_{j,t}$ was included, which equals 1 if policy measure $j$ was in effect on day $t$, and 0 otherwise. The parameter $\gamma_j$ can be interpreted as the log expected change in $R_t$ when policy measure $j$ was implemented, conditional on the day of the week, mobility, and the other policy measures. The explanatory variable $\bar{m}_t$ is the sample average of the daily travel distance in the study population on day $t$. A logarithmic transformation of $m_t$ was used to measure changes in mobility on a relative scale. The parameter $\theta$ is the main parameter of interest and captures the effect of mobility. It can be interpreted as the expected percentage change in $R_t$ associated with a 1 % change in mobility, conditional on the day of the week and the policy measures.

A property of the gamma distribution is that the variance is proportional to the mean. Specifically, if the effective reproductive number is gamma distributed, its variance function on day $t$ is given by

$$\frac{\alpha}{\beta_t^2} = \underbrace{\left(\frac{\alpha}{\beta_t}\right)^2}_{(\mu(R_t))^2} \frac{1}{\alpha} = \frac{(\mu(R_t))^2}{\alpha}. \tag{B.3}$$

This shows that the estimated variance of $R_t$ on a given day is proportional to the square of the expected value predicted by the regression function. The property enables us to capture that $R_t$ may vary more, and, hence, be more uncertain as it takes larger values.

*B.3. Models for subgroups*

We further analysed how the effect of mobility by mode of transport, travel purpose, and sociodemographic subgroup (age group, household size, employment status, and urbanization level) differed between the waves. Here, we used a separate model for each mode of transport, travel purpose, and sociodemographic subgroup. The respective average daily travel distance was used as explanatory variable, i. e. the regression function was defined as

$$\mu(R_t) = \exp(\delta_{\mathrm{wd}(t)} + \sum_{j=1}^{J} \gamma_j p_{j,t} + \theta \log \bar{m}_{k,t}) \ , \tag{B.4}$$

where $\bar{m}_{k,t}$ denotes the average daily travel distance for mode of transport, travel purpose, or sociodemographic subgroup $k$. Apart from the changed explanatory variable, the model specification was the same as for the main model.

*B.4. Models for movement radius*

We also analysed the effects of the movement radius. Five different categories for the movement radius were distinguished, i. e., residential (<500 m), local (500 m–2 km), municipal (2 km–10 km), regional (10–50 km), and long-range (>50 km). Here, the model used was similar to the main model, but, instead of the average travel distance, the share of the population that travelled within a certain movement radius was defined as explanatory variable. The model is

$$\mu(R_t) = \exp(\delta_{\mathrm{wd}(t)} + \sum_{j=1}^{J} \gamma_j p_{j,t} + \theta_l d_{l,t}) \ , \tag{B.5}$$

where $d_{l,t}$ is the share of the population travelling within movement radius $l$ on day $t$. Note that, in this model, no logarithmic transformation of mobility was used as the share is already on a relative scale. Thus, $\theta_l$ can be interpreted as the expected percentage change in $R_t$ associated with an increase in the share of the population travelling in movement radius $l$ by one percentage point, conditional on the day of the week and the policy measures. A separate model was fitted for each movement radius.

*B.5. Model priors*

Table B.2 provides an overview over the choices of priors for the parameters in our models. We used weakly informative priors for all parameters. The priors on the parameters related to policy measures and mobility reflect weak assumptions about the range of realistic effect sizes. All other priors were informed by the general recommendations from the Stan development team.[6]

Similar to the approach taken by Brauner et al.,[4] we modelled the effects of policy measures using an asymmetric Laplace prior on the parameters $\gamma_j$. This prior allows for both positive and negative effects of policy measures since it cannot be a priori excluded that policy measures may lead to increases in the effective reproduction number. With the prior chosen, 90 % of the probability mass was placed on positive effects, implying that policy measures were assumed to be far more likely to reduce the effective reproduction number than to increase it. The highest probability was assigned to an effect size of zero. The scale parameter of the asymmetric Laplace distribution furthermore reflects the assumption that policy measures may reduce $R_t$ by up to 50 %. Note that we modelled the effects of policy measures *conditional* on mobility. The conditional effects are most likely smaller than the unconditional effects of policy measures. The parameter for the effect of mobility was also given an asymmetric Laplace prior. Again, both positive and negative effects were allowed. This implies that decreases in mobility may also lead to increases in the effective reproduction number, but 90 % of the probability mass was placed on positive effects and the highest probability was assigned to an effect size of zero. The scale parameter was chosen such that for a 1 % decrease in mobility, a 1 % reduction in the effective reproduction number is expected, and reductions of up to 3 % are probable. In the models for the different modes of transport, travel purposes, and sociodemographic subgroups, the parameter for the effect of mobility is given the same prior as in the main model.

The effects of movement radius were given a normal prior centred at zero, since it cannot be a priori known whether the increase in the share of a certain movement radius would increase or decrease the effective reproduction number. The standard deviation chosen reflects our expectation that, for a one percentage point increase in the share of a movement radius, an increase or decrease in $R_t$ of more than 20 % is unlikely.

The priors on the weekday-specific intercepts are weakly informative and encompass potential relative differences between weekdays due to reporting and mobility patterns. The shape parameter $\alpha$ of the gamma distribution for $R_t$ was also given a weakly informative prior. The rate parameter $\beta_t$ of the gamma distribution for $R_t$ is however not assigned a prior since it is calculated as $\frac{\alpha}{\mu(R_t)}$. Therefore its prior is implicitly given by the priors on $\alpha$ and $\mu(R_t)$. Similarly, the variance of $R_t$ is a function of $\mu(R_t)$ and $\alpha$, hence its prior is also defined implicitly.

| Parameter | Description | Prior | Models |
|---|---|---|---|
| $\theta$ | Mobility effect | ALD($\lambda = 3, \kappa = 0.35$) | (I) |
| $\theta_k$ | Mobility effect for mode, purpose, or subgroup $k$ | ALD($\lambda = 3, \kappa = 0.35$) | (II) |
| $\theta_l$ | Mobility effect for movement radius $l$ | Normal($\mu = 0, \sigma = 10$) | (III) |
| $\gamma_j$ | Policy measure effect | ALD($\lambda = 15, \kappa = 3$) | (I), (II), (III) |
| $\delta_{\text{wd}(t)}$ | Intercept for weekday wd($t$) | Student-t($\nu = 3, \mu = 0, \sigma = 1$) | (I), (II), (III) |
| $\alpha$ | Shape parameter of gamma distribution for $R_t$ | Exponential(0.05) | (I), (II), (III) |

ALD: asymmetric Laplace distribution

**Table B.2: Prior choices for model parameters.** (I) refers to the main model, i. e., for the total average daily distance. (II) refers to the models for modes of transport, travel purposes, or sociodemographic subgroups. (III) refers to the model for movement radius.

### C. Model diagnostics

The parameter estimates were analysed with common Bayesian model diagnostics to ensure model fit, a sufficient effective sample size, convergence of the chains, and absence of highly influential observations. Here, we followed recommendations by Gelman et al.[7] and the Stan development team[8].

#### *C.1. Posterior predictive checks*

Model fit was assessed by comparing the posterior predictive distribution with the given estimates of the effective reproduction number. For each model, we simulated 4000 draws from the posterior distribution of the model parameters to predict $R_t$ values for the same time periods used to fit the model and compared them to the given $R_t$ estimates (Fig. C.7). The posterior predictive distributions fitted the given estimates well, indicating good fit of all models. For brevity, we only show the fit for the main model in Fig. C.7, the posterior predictions from all other models were similar.

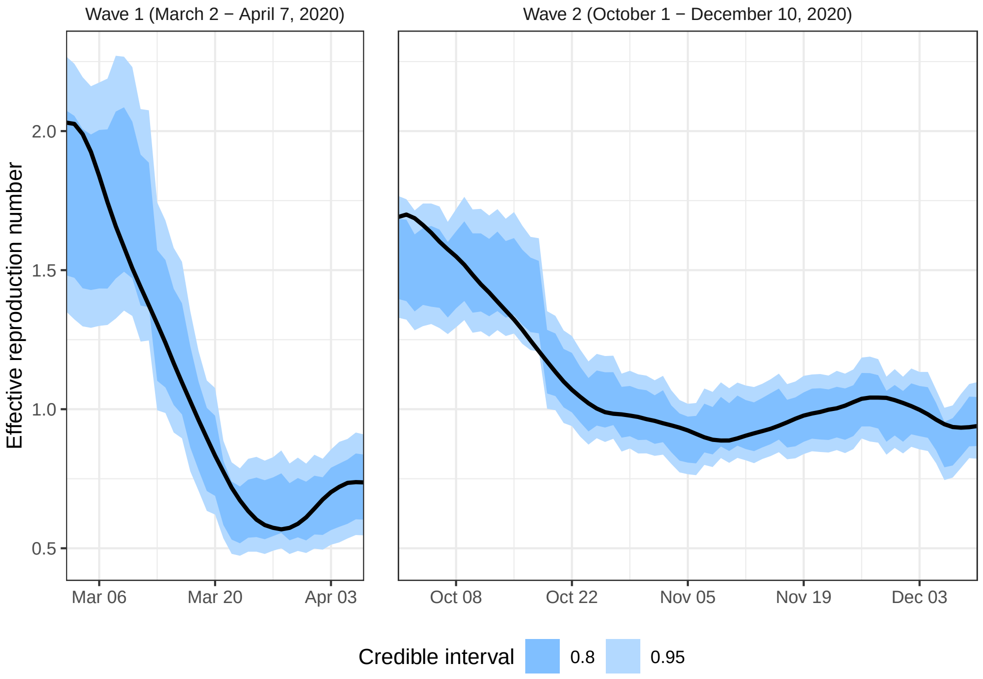


**Figure C.7: Comparison of posterior predictive distribution with observed values.** The black line shows the given estimates of $R_t$. Shaded areas (dark and light blue) represent the 80 % and 95 % credible intervals of model predictions, respectively.

### *C.2. Effective sample size*

The ratio of the effective sample size to the total sample size[8] was computed for each model parameter (Fig. C.8). The ratio was around 0.5 for most parameters and above 0.1 for all parameters, indicating a sufficient number of independent draws from the posterior distribution.

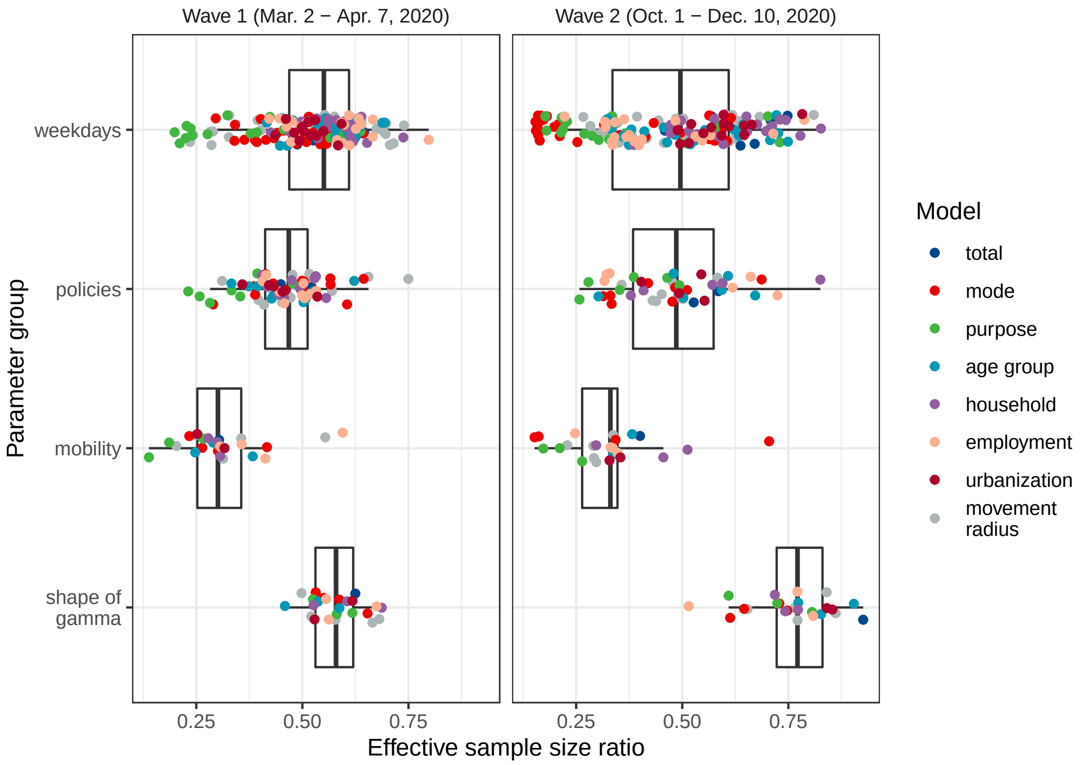


**Figure C.8: Ratio of the effective sample size to the total sample size.** Dots and boxplots show the ratios of the effective sample size to the total sample size for different parameter groups and models in both waves. Values below 0.1 (i. e., fewer than 400 effective samples out of 4000), which are generally considered problematic, did not occur in any model.

### *C.3. Convergence*

For each model parameter, convergence of the Markov chains was assessed via the Gelman-Rubin convergence diagnostic $\hat{R}$.[7] All $\hat{R}$ values are below 1.01, indicating convergence[9] (Fig. C.9).

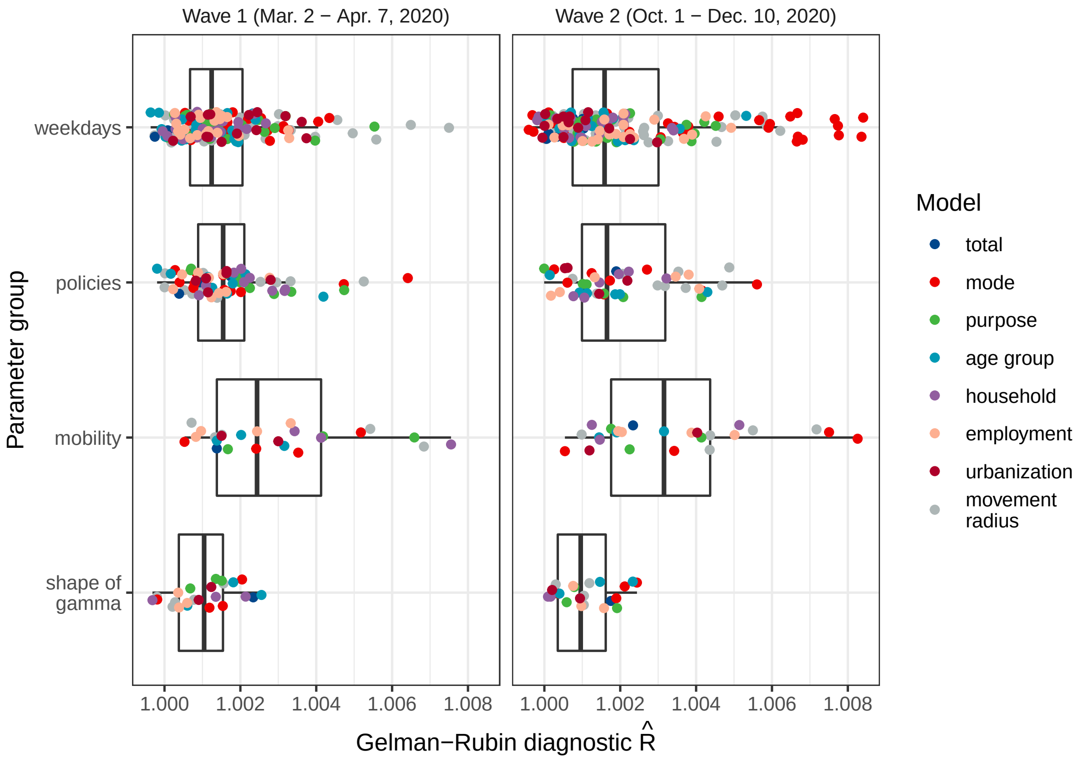


**Figure C.9: Gelman-Rubin convergence diagnostic ($\hat{R}$).** Dots and boxplots show the $\hat{R}$ values for different parameter groups and models in both waves. Values above 1.01 are considered as an indicator of convergence problems[7] but did not occur for any model parameter.

#### *C.4. Influential observations*

We checked for influential observations in the data using the tail shape parameter $k$ of the Pareto importance distribution of each observation, obtained from approximate leave-one-out (LOO) cross-validation with Pareto-smoothed importance sampling (PSIS). The R package loo, version 2.3.1, was used to estimate the Pareto tail shape parameter. Observations with a value of $k$ less than 0.5 are usually considered not influential, and observations with a value between 0.5–0.7 are considered slightly influential.[10, 11] No model had observations with an estimated value of $k$ above 0.7 (Fig. C.10), suggesting absence of highly influential observations.

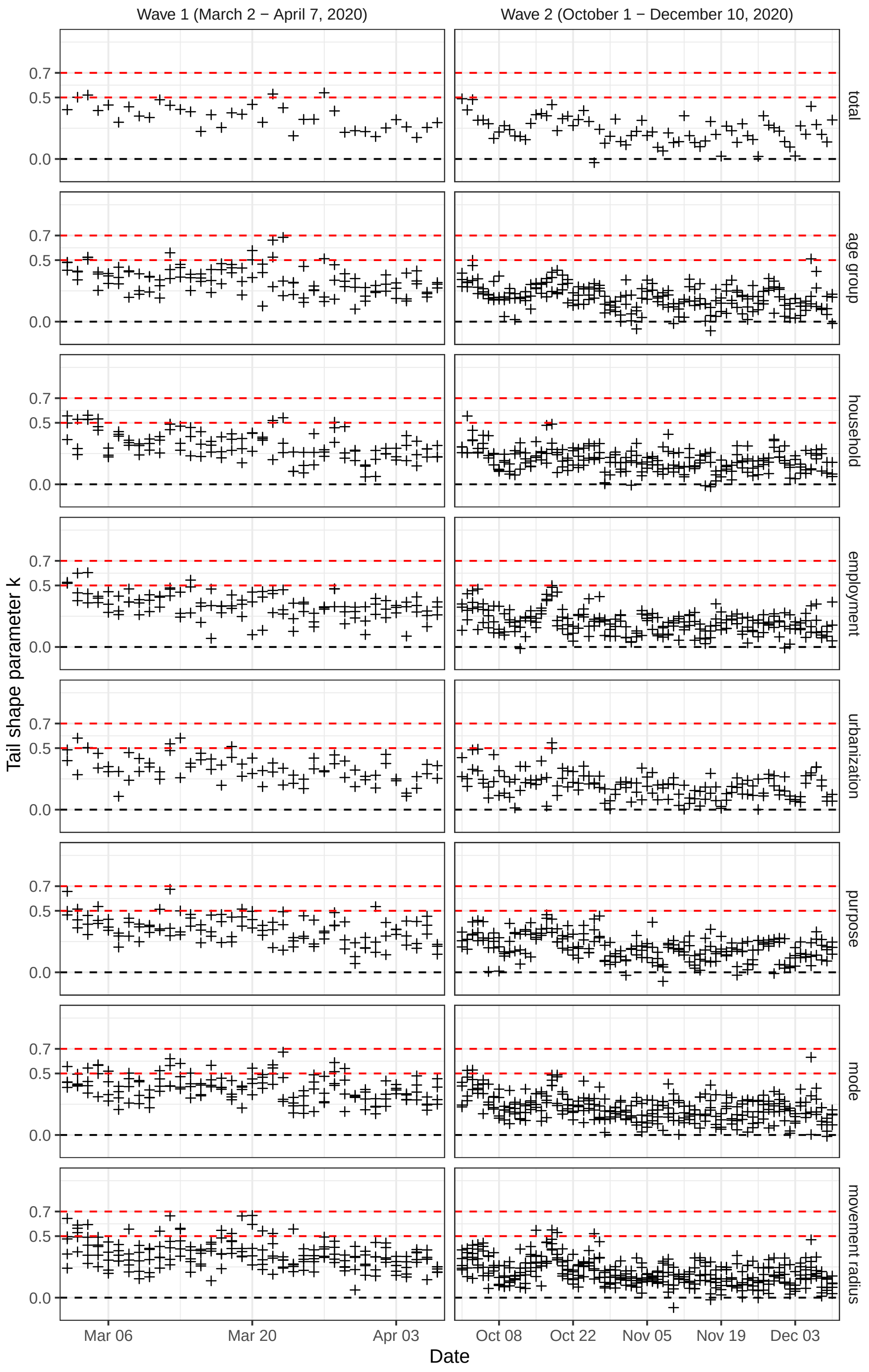


**Figure C.10: PSIS-based check for influential observations.** Crosses show the values of the tail shape parameter $k$ of the Pareto importance distribution of each observation, for each model and wave. Note that all models of a certain category are summarized in one pane. Observations with values below 0.5 are considered not influential, observations with a value between 0.5–0.7 are considered slightly influential, hence the present estimates are not driven by highly influential observations.

### D. Robustness checks

The robustness of the estimated mobility effects was analysed with respect to alternative study periods, alternative estimates of the effective reproduction number, and an alternative model specification with autocorrelated errors. Furthermore, the model was extended to incorporate changes in testing intensity.

#### *D.1. Study periods*

We checked the robustness of our results regarding the specific choice of study periods. In our study, we analysed the time periods from March 2–April 7, 2020 (wave 1) and October 1–December 10, 2020 (wave 2). To assess potential sensitivity to this choice, we also tested how the effect estimates changed when slightly extending or shortening the study periods.

As shown in Fig. D.11, the differences in estimates from alternative study periods were small. The estimates for study periods with a start date one week later were slightly smaller. This finding was expected because, in both waves, the first weeks of the study periods show the effect of mobility reductions when policy measures were not yet implemented, so their exclusion should slightly reduce the estimated effect of mobility. The estimates for study periods with an end date one week earlier have slightly wider credible intervals, suggesting that the variation of mobility under policy measures observed at the end of the study periods was informative for our estimates.

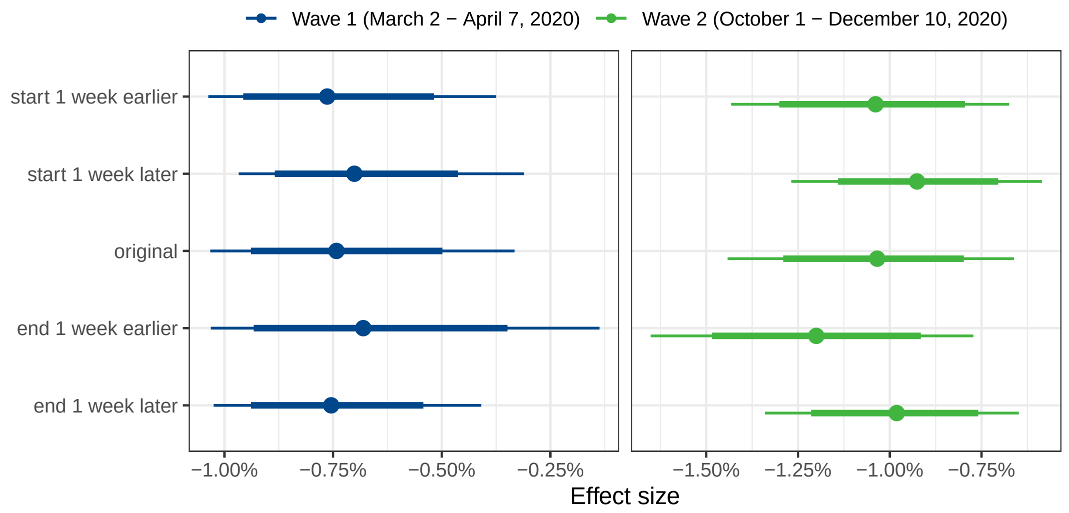


**Figure D.11: Mobility effects for different study periods.** Shown are the estimated changes in $R_t$ given a 1 % reduction in mobility for the original study period and for study periods with either the start or end date shortened or extended by one week. Posterior means (dots) and the 80 % and 95 % credible intervals (thick and thin bars) are reported.

We also estimated effects of mobility for the time between the first and second wave. It is important to note that, for the time period in between the waves, estimates of the effective reproduction number were highly uncertain. This can be explained by the overall small number of reported cases. Moreover, the proportion of infectious persons was likely lower than during the waves, as demonstrated by a lower test positivity rate with a minimum of 0·2 % (June 18, 2020) over the summer period. We fitted a separate model for each month between the first and second wave. No significant relationship between mobility and $R_t$ was found for any of the six months between the waves (Fig. D.12).

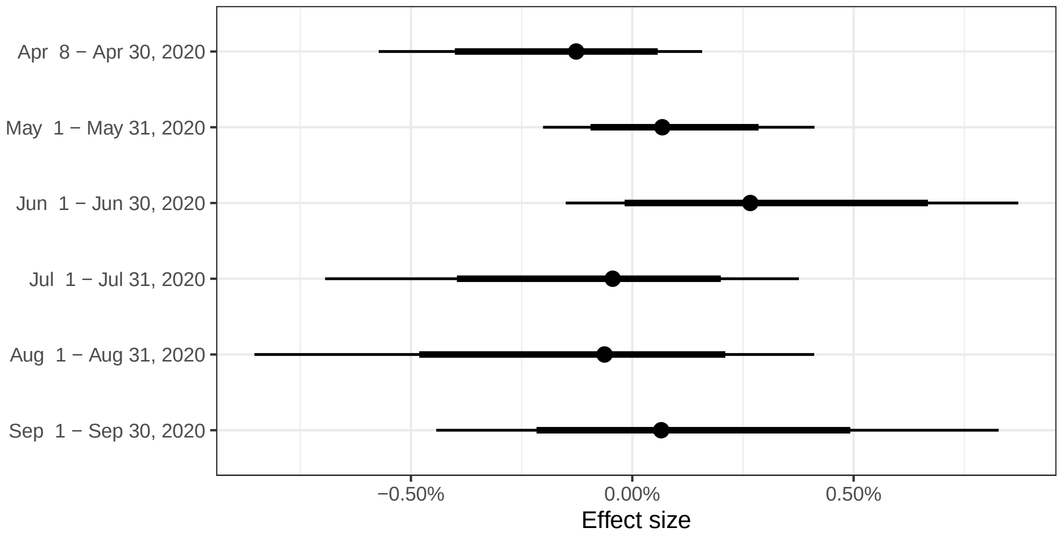


**Figure D.12: Mobility effects for the time period inbetween the first and second wave.** Shown are the estimated changes in $R_t$ given a 1 % reduction in mobility for different months inbetween the first and second wave. Posterior means (dots) and the 80 % and 95 % credible intervals (thick and thin bars) are reported.

*D.2. Other estimates of the effective reproduction number*

In our study, estimates of the effective reproduction number $R_t$ was taken from the Swiss National COVID-19 Science Task Force.[23, 24] Specifically, we used the estimates which were computed from the time series of newly hospitalized patients. The Swiss National COVID-19 Science Task Force also provides alternative estimates based on the time series of (1) confirmed cases or (2) reported deaths. Using these estimates for $R_t$, we present estimates for the effect of mobility in the following.

The estimated mobility effects show some sensitivity to the choice of $R_t$ (Fig. D.13). (1) Compared to estimates based on the number of newly hospitalized patients, estimates based on the number of new confirmed cases yield a smaller estimated effect of mobility in the first wave and a larger estimated effect in the second wave. However, the resulting credible intervals are wider. Generally, estimates based on the number of new confirmed cases are considered potentially sensitive to changes in testing intensity. (2) Estimates based on the number of reported deaths could not be reliably obtained for the beginning of the epidemic and were thus not available for the full study period of the first wave. We therefore report estimates based on the time period March 8 – April 7, 2020 for reported deaths in the first wave. The estimate is smaller than the one based on newly hospitalized patients and again has a wider credible interval. This may be partly due to the missing observations at the beginning of the study period. In the second wave, estimates based on the number of reported deaths yield an estimated mobility effect similar to that based on the number of newly hospitalized patients.

The 80 % credible intervals of effect sizes overlap for all $R_t$ estimates. We thus consider our qualitative findings to be robust to alternative definitions of $R_t$.

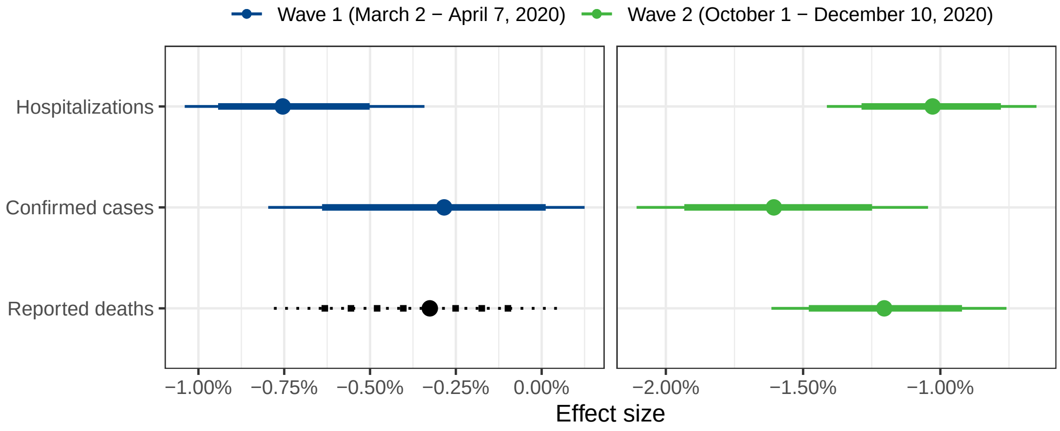


**Figure D.13: Mobility effects for different estimates of $R_t$.** Shown are the estimated changes, given a 1 % reduction in mobility, in different estimates of $R_t$ either based on hospitalized patients, confirmed cases, or reported deaths. The estimate for reported deaths in the first wave was obtained only for observations after March 8, 2020 onward due to limited data availability. Posterior means (dots) and the 80 % and 95 % credible intervals (thick and thin bars) are reported.

#### *D.3. Autocorrelated errors*

To assess the robustness of our main estimates against autocorrelated model errors, which may be present in time series regressions, we used the same regression function for the main model,

$$\mu(R_t) = \exp(\delta_{\text{wd}(t)} + \sum_{j=1}^{J} \gamma_j p_{j,t} + \theta \log \bar{m}_t), \quad \text{(D.1)}$$

and added multiplicative errors $\exp(\epsilon_t)$, where $\epsilon_t = (\frac{R_t - \mu(R_t)}{\sqrt{\widehat{\text{Var}}[R_t]}})$ are Pearson residuals. We model the residuals to follow an AR(2) process, i. e.,

$$\epsilon_t = \tau_1 \epsilon_{t-1} + \tau_2 \epsilon_{t-2} + u_t, \quad \text{(D.2)}$$

The lag order of 2 was chosen by inspecting the autocorrelation function of the posterior predictive errors from the original model. The autocorrelation parameters $\tau_1$ and $\tau_2$ were both given a regularizing normal prior, i. e., $\tau_1, \tau_2 \sim \text{Normal}(0, 0.4)$. The term $u_t$ is the so-called innovation of the AR process and is implicitly defined for any given day $t$ through the other parameters of the model and the residuals of previous days. The specification of the model was otherwise analogous to the main model.

Figure D.14 shows the estimated effects of mobility for the main model without autocorrelated errors and for the model with autocorrelated errors. The model with autocorrelated errors estimates slightly stronger effects of mobility, and the estimates are slightly more uncertain for the second wave. Nevertheless, the qualitative findings remain the same.

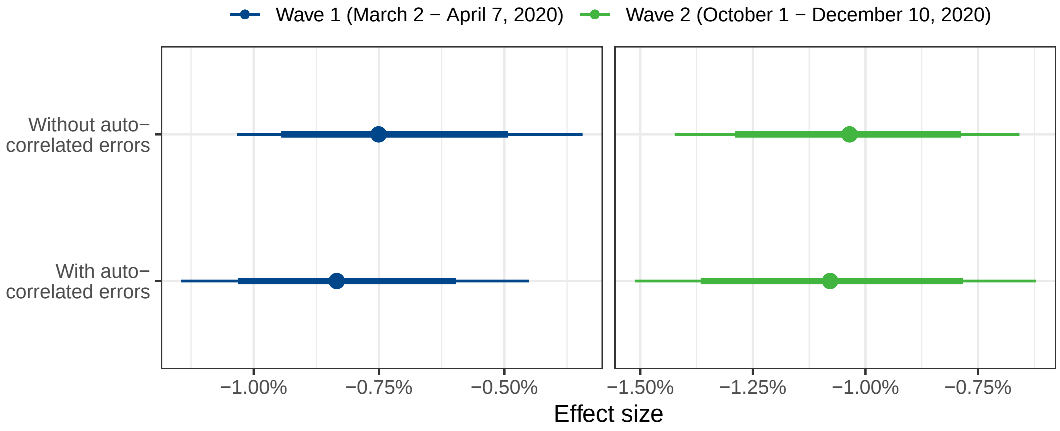


**Figure D.14: Mobility effects without (as in the main paper) and with autocorrelated errors.** Shown are the estimated changes in $R_t$ given a 1 % reduction in mobility, for models without and with autocorrelated errors. Posterior means (dots) and the 80 % and 95 % credible intervals (thick and thin bars) are reported.

#### *D.4. Testing intensity*

Changes in testing intensity over time may influence the effective reproduction number or estimates thereof. On the one hand, an abrupt surge in testing may distort estimates of the effective reproduction number when these are based on confirmed cases, because more testing can lead to a higher proportion of infections that are detected. On the other hand, as detected cases potentially lead to contact tracing and isolation of infectious individuals, a high testing intensity also has the potential to reduce the number of future infections and thus lower the effective reproduction number.

In our study, the direct influence of changes in testing intensity is mostly avoided through the use of $R_t$ estimates that are inferred from the number of newly hospitalized patients. Regarding the indirect effect of testing intensity, we wanted to assess whether testing may confound the effect of mobility in our model. We thus extended the model by adding the share of positive PCR tests for COVID-19 as another predictor. This share, also called test positivity rate, often serves as an indicator of the intensity of testing relative to the current number of confirmed cases. A low positive rate means that many tests are performed in comparison to the number of new cases detected; therefore, testing is rather comprehensive. On the contrary, a high positive test rate may indicate many further cases which have not been detected and confirmed. Similar to our modelling of mobility effects, we smoothed the test positivity rate with a 3-day moving average and applied a logarithmic transformation to measure changes in testing intensity on a relative scale. We furthermore added a lag of 6 days, because tests performed on a given day can only avoid subsequent infections through contact tracing and isolation. Hence, a change in testing should only change the effective reproduction number with a delay of several days. The chosen lag of 6 days roughly equals the mean generation time of COVID-19.[12, 13] We used a broad gamma prior for the effect of testing intensity, here Gamma($\mu = 1, \sigma = 0.5$). The prior reflects the assumption that for a 1 % decrease in the positive rate (which translates to an increase in testing intensity), the effective reproduction number can decrease by up to 6% but will not increase. All other priors are chosen as in the main model.

As shown in Fig. D.15, the estimates for the effect of mobility on the effective reproduction number are almost identical for the models with and without the effect of testing intensity. We thus found no evidence that testing intensity confounded our results.

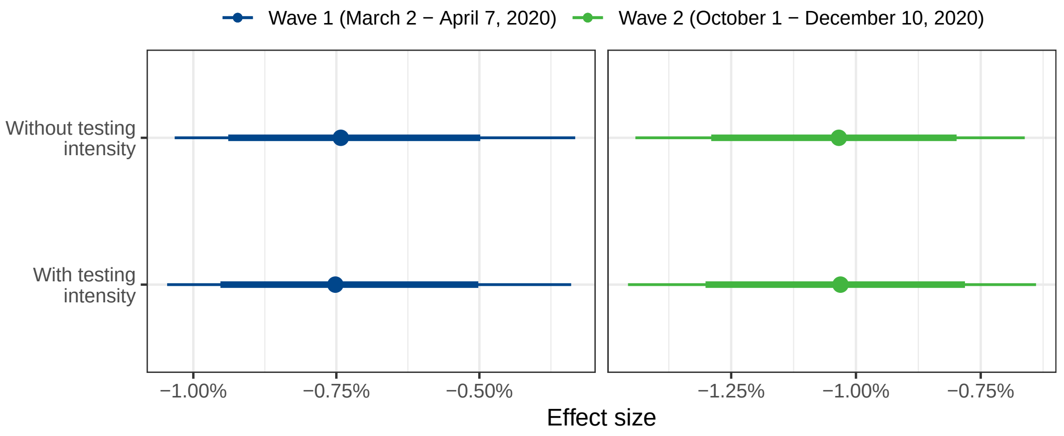


**Figure D.15: Mobility effects without (as in the main paper) and with adjusting for testing intensity.** Shown are the estimated changes in $R_t$ given a 1 % reduction in mobility, without and with adjusting for the effect of testing on further transmission. Posterior means (dots) and the 80 % and 95 % credible intervals (thick and thin bars) are reported.

[Supp4] Brauner JM, Mindermann S, Sharma M, Johnston D, Salvatier J, Gavenčiak T, et al. Inferring the effectiveness of government interventions against COVID-19. *Science*. doi:10.1126/science.abd9338.

[Supp5] Haug N, Geyrhofer L, Londei A, Dervic E, Desvars-Larrive A, Loreto V, et al. Ranking the effectiveness of worldwide COVID-19 government interventions. *Nature Human Behaviour* 1303–1312. doi:10.1038/s41562-020-01009-0.
URL https://www.nature.com/articles/s41562-020-01009-0

[Supp6] Stan development team. Prior choice recommendations. 2020.
URL https://github.com/stan-dev/stan/wiki/Prior-Choice-Recommendations

[Supp7] Gelman A, Rubin DB, et al. Inference from iterative simulation using multiple sequences. *Statistical Science* 1992; **7** (4): 457–472.

[Supp8] Stan development team. Stan language reference manual (version 2.25). 2019.
URL https://mc-stan.org/docs/2_21/reference-manual

[Supp9] Gelman A, Carlin JB, Stern HS, Dunson DB, Vehtari A, and Rubin DB, Bayesian Data Analysis, Chapman & Hall / CRC Texts in Statistical Science, 2013.

[Supp10] Vehtari A, Gelman A, and Gabry J. Practical Bayesian model evaluation using leave-one-out cross-validation and WAIC. *Statistics and Computing* 2017; **27** (5): 1413–1432. doi:10.1007/s11222-016-9696-4.

[Supp11] Vehtari A, Simpson D, Gelman A, Yao Y, and Gabry J. Pareto smoothed importance sampling. *arXiv.*
URL `https://arxiv.org/pdf/1507.02646`

[Supp12] Ganyani T, Kremer C, Chen D, Torneri A, Faes C, Wallinga J, et al. Estimating the generation interval for coronavirus disease (COVID-19) based on symptom onset data, march 2020. *Euro surveillance: European communicable disease bulletin* 2020; **25** (17):. `doi:10.2807/1560-7917.ES.2020.25.17.2000257`.

[Supp13] Ferretti L, Ledda A, Wymant C, Zhao L, Ledda V, Abeler-Dörner L, et al. The timing of COVID-19 transmission. *medRxiv.* `doi:10.1101/2020.09.04.20188516`.

Strengthening the Reporting of Observational Studies in Epidemiology (STROBE) Statement

| Section and item | Item No | Recommendation | Reported on page |
|---|---|---|---|
| **Title and abstract** | 1 | (a) Indicate the study's design with a commonly used term in the title or the abstract | 1 |
| | | (b) Provide in the abstract an informative and balanced summary of what was done and what was found | 1 |
| **Introduction** | | | |
| Background/rationale | 2 | Explain the scientific background and rationale for the investigation being reported | 1–2 |
| Objectives | 3 | State specific objectives, including any prespecified hypotheses | 2 |
| **Methods** | | | |
| Study design | 4 | Present key elements of study design early in the paper | 2–3 |
| Setting | 5 | Describe the setting, locations, and relevant dates, including periods of recruitment, exposure, follow-up, and data collection | 3–4 |
| Participants | 6 | (a) Give the eligibility criteria, and the sources and methods of selection of participants | NA |
| Variables | 7 | Clearly define all outcomes, exposures, predictors, potential confounders, and effect modifiers. Give diagnostic criteria, if applicable | 4–5 |
| Data sources / measurement | 8 | For each variable of interest, give sources of data and details of methods of assessment (measurement). Describe comparability of assessment methods if there is more than one group | 3–4 + Supp. 1–2 |
| Bias | 9 | Describe any efforts to address potential sources of bias | NA |
| Study size | 10 | Explain how the study size was arrived at | 3 |
| Quantitative variables | 11 | Explain how quantitative variables were handled in the analyses. If applicable, describe which groupings were chosen and why | 4–5 |
| Statistical methods | 12 | (a) Describe all statistical methods, including those used to control for confounding | 4–5 + Supp. 3–11 |
| | | (b) Describe any methods used to examine subgroups and interactions | 4–5 + Supp. 5 |
| | | (c) Explain how missing data were addressed | NA |
| | | (d) If applicable, describe analytical methods taking account of sampling strategy | NA |
| | | (e) Describe any sensitivity analyses | Supp. 12–17 |
| **Results** | | | |
| Participants | 13 | (a) Report numbers of individuals at each stage of study–eg numbers potentially eligible, examined for eligibility, confirmed eligible, included in the study, completing follow-up, and analysed | NA |
| | | (b) Give reasons for non-participation at each stage | NA |
| | | (c) Consider use of a flow diagram | NA |
| Descriptive data | 14 | (a) Give characteristics of study participants (eg demographic, clinical, social) and information on exposures and potential confounders | 6–9 + Supp. 1–2 |
| | | (b) Indicate number of participants with missing data for each variable of interest | NA |
| Outcome data | 15 | Report numbers of outcome events or summary measures | |
| Main results | 16 | (a) Give unadjusted estimates and, if applicable, confounder-adjusted estimates and their precision (eg, 95% confidence interval). Make clear which confounders were adjusted for and why they were included | 10–13 |
| | | (b) Report category boundaries when continuous variables were categorized | NA |
| | | (c) If relevant, consider translating estimates of relative risk into absolute risk for a meaningful time period | NA |
| Other analysis | 17 | Report other analyses done—eg analyses of subgroups and interactions, and sensitivity analyses | 10–13 + Supp. 12–17 |
| **Discussion** | | | |
| Key results | 18 | Summarise key results with reference to study objectives | 13–14 |
| Limitations | 19 | Discuss limitations of the study, taking into account sources of potential bias or imprecision. Discuss both direction and magnitude of any potential bias | 14–15 |
| Interpretation | 20 | Give a cautious overall interpretation of results considering objectives, limitations, multiplicity of analyses, results from similar studies, and other relevant evidence | 13–15 |
| Generalisability | 21 | Discuss the generalisability (external validity) of the study results | 14 |
| **Other information** | | | |
| Funding | 22 | Give the source of funding and the role of the funders for the present study and, if applicable, for the original study on which the present article is based | 20 |
